# Care Models for the Genetic Evaluation of Dilated Cardiomyopathy at Sites of the DCM Consortium

**DOI:** 10.64898/2026.04.06.26350275

**Authors:** Elizabeth Jordan, Tia Moscarello, Hibatallah Khafagy, Patricia Parker, Phoenix Grover, Simone Weinmann, Joseph Liu, Alberta Nomo, Naomi Barker, Emily E Brown, Akos Berthold, Jessica Chowns, Susan Christian, Amy Ekwurtzel, Judy Fan, Monisha Kisling, Daria Ma, Erin Miller, Jessica Sweeney, Brian Reys, Nancy Robles, Lisa Von Wald, Wendy Flowers, Greg Hershberger, Krishna Aragam, Michael A. Burke, Jamie Diamond, Mark H. Drazner, Gregory A. Ewald, Stephen S. Gottlieb, Garrie Haas, Mark Hofmeyer, Gordon S. Huggins, Javier Jimenez, Daniel P. Judge, Stuart Katz, Masataka Kawana, Evan P. Kransdorf, Cindy M. Martin, Elina Minami, Anjali Owens, Palak Shah, Chetan Shenoy, Supriya Shore, Frank Smart, Douglas Stoller, Jose Tallaj, W. H. Wilson Tang, Jessica Wang, Jane Wilcox, Ray E. Hershberger

## Abstract

**Background:** Clinical genetic evaluation for patients with dilated cardiomyopathy (DCM) is minimally implemented and models of care are not defined. To understand current genetics care for DCM, a systematic needs assessment was conducted.

**Methods:** Principal Investigators (PIs) of the DCM Consortium convened at the Summer Scientific Symposium in July 2025. An electronic needs assessment was collected from the 24 PIs in advance to define current care models by evaluating which Heart Failure Society of America-recommended genetic evaluation components are conducted, by whom, and time required. Descriptive statistics were generated to characterize model features. Focus group discussions explored barriers and facilitators to implementing genetic services.

**Results:** Four care models emerged from the PI responses: 1 – *Traditional-Synchronous* (25%, n=6, requiring the most time per patient), 2 – *Traditional-Asynchronous* (33%, n=8), 3 – *Externally Sourced* (17%, n=4), and 4 – *Physician/Advanced Practice Provider Conducted* (25%, n=6, requiring the least time per patient). All models used genetic testing, whereas other components were implemented variably or not at all. Models 1 (15.7*±*4.1) and 2 (15.4*±*3.0) were rated more acceptable than Model 4 (9.8*±*2.9; 1 vs 4: *p*=0.027; 2 vs 4, *p*=0.023). Notably, 88% of PIs used genetic information for treatment decisions, including ICD placement (83%; n=20) or cardiac transplant (63%; n=15). Major facilitator themes from focus group discussions included having a genetic counselor on the HF team and developing authoritative standards directing provision of DCM genetic services. Barrier themes included operational challenges, limited personnel, clinician under-recognition, need for new service delivery models, and billing/reimbursement.

**Conclusions:** DCM genetic care models and components were highly variable across the 24 sites of the DCM Consortium, even though all sites discussed similar factors that enable or hinder implementing genetic services for DCM. Understanding the basis of practice model variability may provide insight to yield more scalable care approaches.

*Clinical Perspective:* What is new? - Four care models to deliver genetic evaluation services for the assessment of dilated cardiomyopathy (DCM) were identified across the 24 sites of the DCM Consortium.
- Despite universal use of genetic testing and shared barriers and facilitators across sites, substantial variability exists across and within identified model types, and clinical genetic evaluation is not routinely implemented for all patients with DCM. What are the clinical implications? - For Heart Failure (HF) centers seeking to implement genetics services for DCM patients and families, the DCM Consortium describes the scaffolding of current care models implemented at HF centers across the US to inform local program development.
- The incompletely and variably implemented care models highlight the need for a standardized, scalable framework to enhance consistent and equitable integration of clinical genetic evaluation into routine DCM care.

## INTRODUCTION

Dilated cardiomyopathy (DCM), a major cause of heart failure (HF), has a substantial genetic basis.^1^ Current scientific statements^2,3^ and guidelines^4–6^ recommend genetic evaluation for patients with DCM, but implementation has been challenging and uptake of genetic testing (GT) has been minimal.^7–9^ Others have addressed the underutilization of clinical genetics practice in cardiomyopathy,^7–10^ and investigations of genetic services for all heritable cardiovascular diseases have demonstrated similar challenges, with barriers including education, cost, and lack of access to genetic counselors (GCs) and/or clinical genetics expertise on the care team.^11,12^

In 2024, the investigators of the DCM Consortium,^13^ a group organized to conduct genetic research in DCM, convened to identify issues and target efforts to improve the clinical implementation of precision medicine for DCM in their US HF clinics. The results of that needs assessment identified major challenges of billing, workforce, education, and managing genetic uncertainty.^14^ To gain further insight into these issues and to consider remedies, in 2025 the DCM Consortium investigators sought to extend their prior work with a systematic investigation of each institution’s current clinical genetics care model for DCM patients at their centers, and to codify each institution’s operational approach. Here investigators of the DCM Consortium provide a survey of the characteristics, strengths, and challenges of their current models, and provide a blueprint for HF clinicians seeking to implement genetic services at their institutions.

## METHODS

The single institutional review board (IRB) for the DCM Research Project at the University of Pennsylvania deemed this assessment exempt.

### Survey Design

A web-based Qualtrics survey was completed by site principal investigators (PIs, all cardiologists) of the DCM Consortium between June 20 to July 11, 2025. There were four sections to the survey: (A) Your/Your Institution’s Information, (B) Genetic Evaluation Practices, (C) Operational Approach to Genetic Evaluation, and (D) Goals for the Future and Final Comments. Section C (Operational Approach to Genetic Evaluation) included the acceptability implementation measure (AIM)^15^ where participants rated four statements regarding their current practice model on a Likert scale from 1 (completely disagree) to 5 (completely agree) to the acceptability of their current model, where “acceptability” is defined as *the perception among implementation stakeholders that a given treatment, service, practice, or innovation is agreeable, palatable or satisfactory.*^16^ The AIM has been previously validated, demonstrating good internal consistency (Cronbach’s α = 0.83) and strong test–retest reliability (attenuation-corrected Pearson correlation = 0.80).^15^ The full survey instrument is provided in the supplement.

### Focus Group Discussion

Open text responses and Section C items (*Operational Approach to Genetic Evaluation*) were analyzed to identify patterns in practice structure and workflows to group similar care approaches in advance of the in-person 2025 Summer Scientific Symposium event. Two investigators (EJ, TM) coded the responses separately to identify themes at each center that distinguished similar patterns that defined model types. Focus group discussions with DCM Consortium site PIs and other site team representatives present at the Symposium were conducted within each group as identified and assigned by the investigators. A structured question guide was used to guide the discussion groups which was facilitated by a site-affiliated GC present at the Symposium. A COREQ checklist^17^ and coding tree are provided in the Supplemental Material.

### Analysis

Descriptive statistics (percentages, medians, means, and standard deviations) were generated. Because Likert scale responses to the AIM^15^ are ordinal and not normally distributed, group differences were assessed using Kruskal-Wallis tests, followed by Dunn’s post-hoc tests with Sidak correction for pairwise comparisons. Statistical significance was defined as p<0.05.

Focus group discussions were recorded, transcribed and analyzed for themes. As defined by the Consolidated Framework for Implementation Research, barriers were considered factors that hinder implementation of DCM genetic services and facilitators as those that enable or support successful integration into care.^18^ Transcripts were coded for themes using a constant comparative method,^19^ a reductive and interpretive method that allows themes representing meaningful patterns of common messages to emerge. Two investigators separately read and analyzed the transcribed data (TM, HK) resulting in two independent sets of codes. The investigators generated independent notes with tentative interpretations of explicit and implicit meanings and comparisons of the content with other sources. The investigators met to compare notes and refine the codebook. Investigators (TM, HK) coded each transcript independently and then met to adjudicate passages and codes. All four transcripts were coded and re-coded with the updated codebook. The final codebook is provided in the Supplemental Material.

## RESULTS

All site principal investigators (PIs; n=24, 100%) participated in the survey. Demographic and training backgrounds of the 24 site PIs have been previously published: most were non-Hispanic white males who were HF/transplant (TX) trained cardiologists based at academic medical centers. Site characteristics included active HF/TX inpatient and outpatient programs, ranging in size from medium to large volumes of patients, as previously presented.^14^

Care models have been defined in Figure 1 and have been further described below. Clinical team members participating in each care model have been summarized in Table 1, with an average of 5.6 HF/TX physicians per site participating in genetic evaluation. 17 (71%) centers had access to GCs internal to their institution, including 6 with cardiovascular-specialized GCs integrated into the HF and/or cardiology team. One site had access to genetics expertise but not within their institution and six did not have access to any genetics professionals. On average 1.6 advanced practice providers (APPs) also participated in genetic evaluation component(s), with 29% of centers having an APP with genetics expertise on their team.

**Figure 1.**
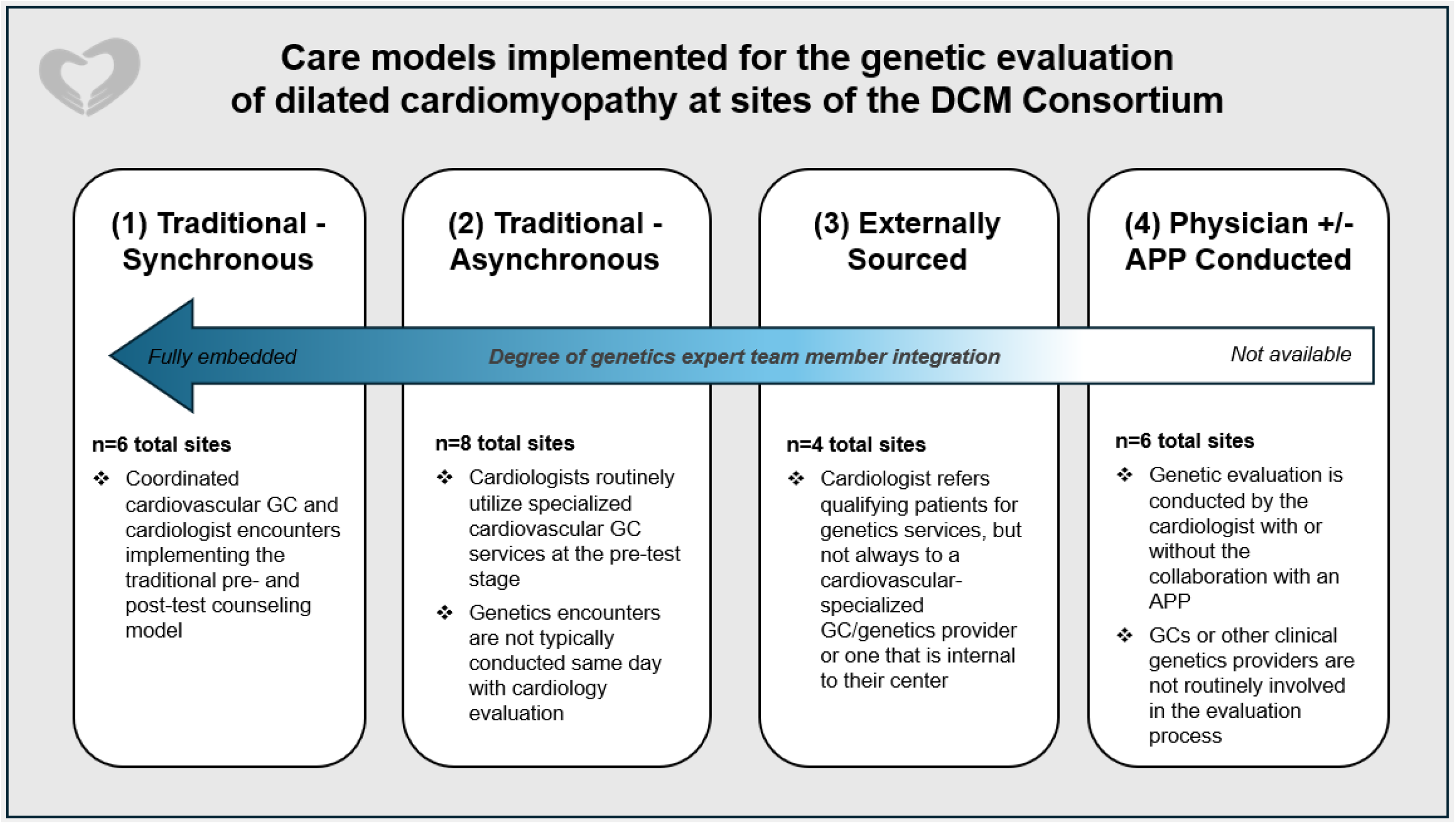
Current care models for genetic evaluation of dilated cardiomyopathy at sites of the DCM Consortium. The arrow points to increasing degrees of genetics expert team member integration into the model of care with darkened color showing higher integration; expert team members include genetic counselors and geneticists. DCM=dilated cardiomyopathy; GC=genetic counselor; pre-test means before genetic testing; APP = advanced practice provider including a certified nurse specialist or nurse practitioner.

**Table 1.**
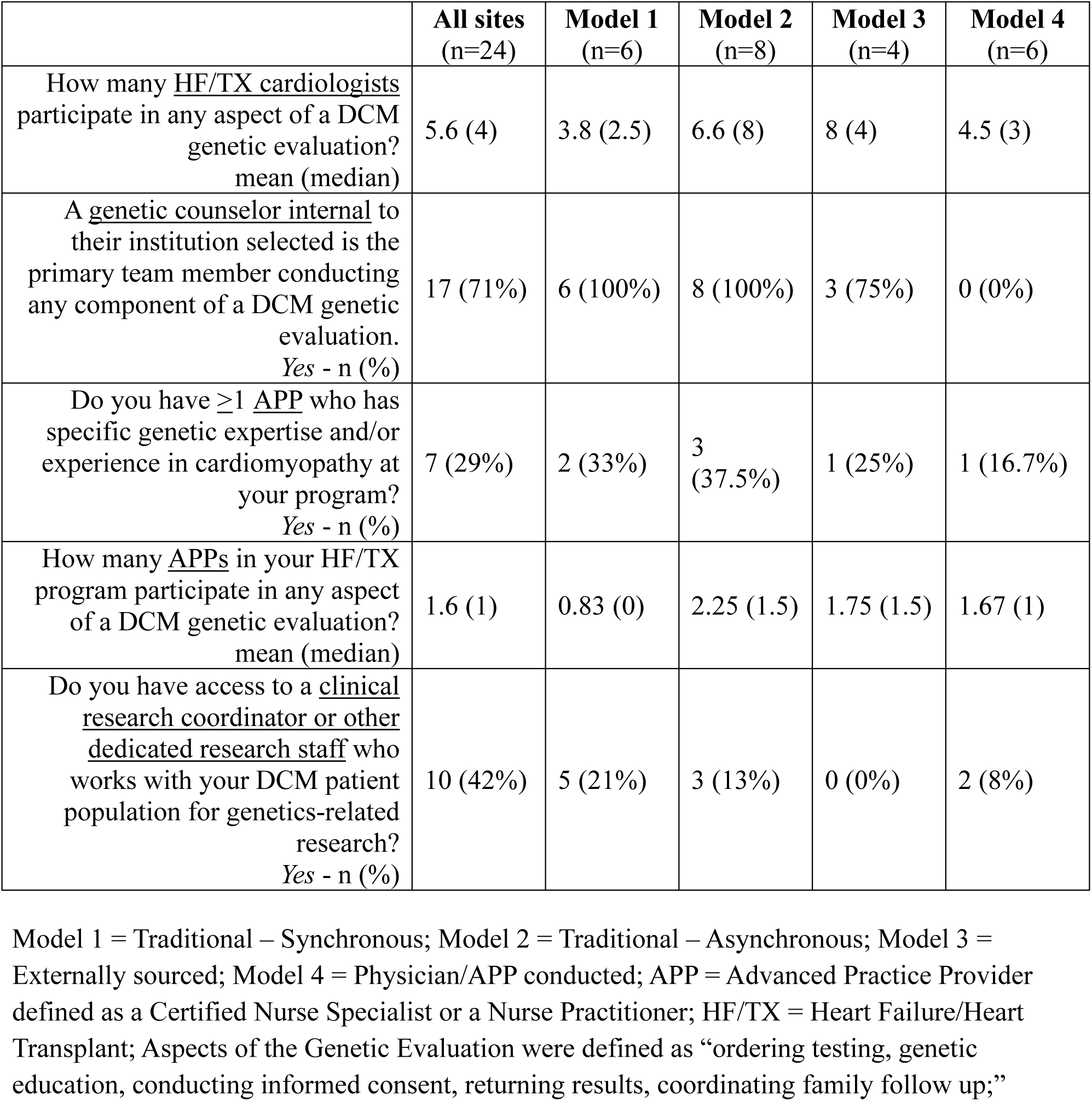
Physician and Advanced Practice Provider Participation in the DCM Genetic Evaluation by Practice Model.

|  | **All sites**<br>**(n=24)** | **Model 1**<br>**(n=6)** | **Model 2**<br>**(n=8)** | **Model 3**<br>**(n=4)** | **Model 4**<br>**(n=6)** |
| --- | --- | --- | --- | --- | --- |
| How many <u>HF/TX cardiologists</u> participate in any aspect of a DCM genetic evaluation?<br>mean (median) | 5.6 (4) | 3.8 (2.5) | 6.6 (8) | 8 (4) | 4.5 (3) |
| A <u>genetic counselor internal</u> to their institution selected is the primary team member conducting any component of a DCM genetic evaluation.<br>Yes - n (%) | 17 (71%) | 6 (100%) | 8 (100%) | 3 (75%) | 0 (0%) |
| Do you have $\geq 1$ <u>APP</u> who has specific genetic expertise and/or experience in cardiomyopathy at your program?<br>Yes - n (%) | 7 (29%) | 2 (33%) | 3 (37.5%) | 1 (25%) | 1 (16.7%) |
| How many <u>APPs</u> in your HF/TX program participate in any aspect of a DCM genetic evaluation?<br>mean (median) | 1.6 (1) | 0.83 (0) | 2.25 (1.5) | 1.75 (1.5) | 1.67 (1) |
| Do you have access to a <u>clinical research coordinator or other dedicated research staff</u> who works with your DCM patient population for genetics-related research?<br>Yes - n (%) | 10 (42%) | 5 (21%) | 3 (13%) | 0 (0%) | 2 (8%) |

Model 1 = Traditional – Synchronous; Model 2 = Traditional – Asynchronous; Model 3 = Externally sourced; Model 4 = Physician/APP conducted; APP = Advanced Practice Provider defined as a Certified Nurse Specialist or a Nurse Practitioner; HF/TX = Heart Failure/Heart Transplant; Aspects of the Genetic Evaluation were defined as “ordering testing, genetic education, conducting informed consent, returning results, coordinating family follow up;”

### Four Care Models for DCM Genetic Evaluation

Thematic analysis of survey responses revealed four models of care, as defined in Figure 1: 1 - *Traditional-Synchronous* (n=6), 2 - *Traditional-Asynchronous* (n=8), 3 - *Externally Sourced* (n=4), and 4 *- Physician +/- APP Conducted* (n=6). Estimates were provided for the time required to implement core components (Figure 2), as defined by the Heart Failure Society of America (HFSA),^5^ and varied widely for each model, with Model 1 requiring the most time per patient (128 minutes) and Model 4 the least (67.9 minutes; Table 2). Composite scores of acceptability using the AIM differed across model types, where Model 1 (15.7 *±* 4.1) and Model 2 (15.4 *±* 3.0) were overall rated significantly more acceptable than Model 4 (9.8 *±* 2.9; Model 1 vs Model 4: *p* = 0.027; Model 2 vs Model 4, *p* = 0.023; Table 3).

**Figure 2.**
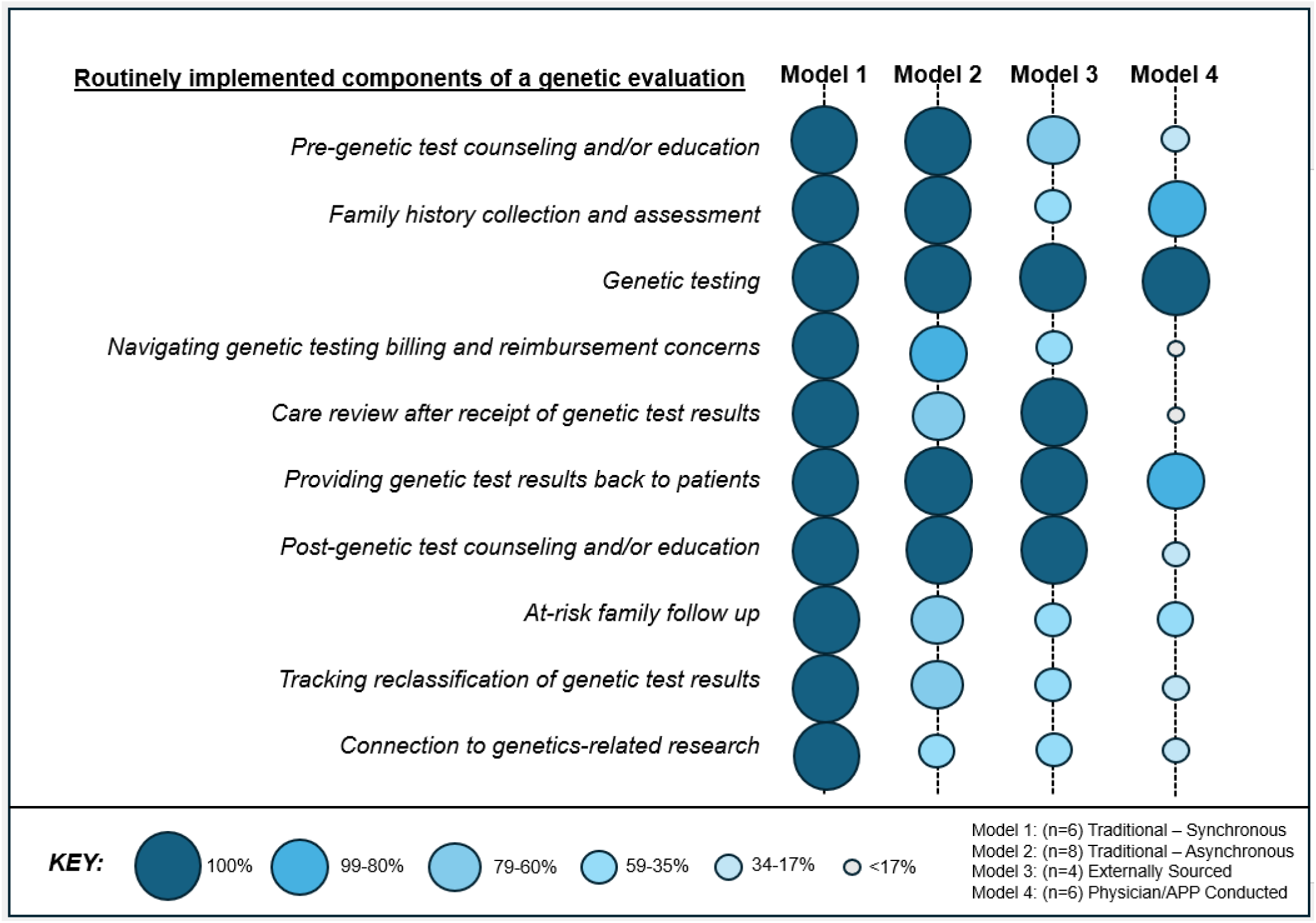
Aspects of a genetic evaluation routinely offered in the currently implemented models for DCM patients at DCM Consortium Sites. The percentage of sites who routinely implement components of a genetic evaluation, specified by models of genetic care, are shown by the size and color of the circles.

**Table 2.** Time required to implement components of a DCM genetic evaluation for a single DCM patient under each model type at DCM Consortium sites.

| **Model Type** | **Estimated Time in Minutes to Complete Evaluation Components for an Individual DCM Patient** |  |  |  |  |  |
| --- | --- | --- | --- | --- | --- | --- |
|  | **Pre-Genetic Test Counseling<sup>a</sup>** | **Family History Assessment** | **Navigating Billing and Reimbursement<sup>a</sup>** | **Result Review** | **Post-Genetic Test Counseling<sup>a</sup>** | **Total time to complete all components<sup>c</sup>** |
| Model 1: Traditional – Synchronous (n=6) | 35.5 | 12.5 | 21.8 | 34.3 | 24.3 | **128.4** |
| Model 2: Traditional – Asynchronous (n=8) | 39.9 | 9 | 16.8<sup>b</sup> | 21.8<sup>b</sup> | 24.1<sup>b</sup> | **111.5** |
| Model 3: Externally Sourced (n=4) | 23<sup>b</sup> | 11<sup>b</sup> | 15.5<sup>b</sup> | 30.5 | 15.5 | **95.5** |
| Model 4: Physician +/- APP Conducted (n=6) | 15.5<sup>b</sup> | 6.4 | N/A | 23<sup>b</sup> | 23<sup>b</sup> | **67.9** |

min=minute; N/A=Not Applicable, as there were no sites in this model group that indicated that they navigate billing and reimbursement for genetic services for DCM patients on a routine basis; <sup>a</sup>responses included options of ≤15, 16-30, 31-60, or >60 minutes; medians for these structured ranges were used to calculate average estimated time for each of these items, respectively. <sup>b</sup>Not all sites in this model group indicated that they routinely perform this service component, therefore the denominator is lower than the total sites in the model group. <sup>c</sup>Total time was calculated as the cumulative average time for each component represented across the table for each respective model.

**Table 3.** Acceptability implementation measure (AIM) of current models of genetic evaluation for DCM at sites of the DCM Consortium.

|  | **Model 1**<br>(M1; n=6) | **Model 2**<br>(M2; n=8) | **Model 3**<br>(M3; n=4) | **Model 4**<br>(M4; n=6) | **Significant Pairwise Differences** |
| --- | --- | --- | --- | --- | --- |
| *Item* | *Mean ± Standard Deviation* |  |  |  |  |
| My institution's current practice model for the genetic evaluation of DCM meets my approval. | 3.8 ± 1.2 | 3.9 ± 0.8 | 2.3 ± 1.3 | 2.8 ± 0.9 | -- |
| My institution's current practice model for the genetic evaluation of DCM is appealing to me. | 4.2 ± 1.1 | 3.9 ± 0.8 | 2.5 ± 1.7 | 2.5 ± 0.8 | -- |
| I like my institution's current practice model for the genetic evaluation of DCM. | 3.8 ± 1.1 | 3.9 ± 0.8 | 2.3 ± 1.3 | 2.3 ± 0.7 | M1 vs M4 ($p=0.026$),<br>M2 vs M3 ($p=0.040$),<br>M2 vs M4 ($p=0.013$) |
| I welcome my institution's current practice model for the genetic evaluation of DCM. | 3.8 ± 1.1 | 3.8 ± 0.8 | 2.3 ± 1.6 | 2.2 ± 0.7 | M1 vs M4 ($p=0.028$),<br>M2 vs M4 ($p=0.026$) |
| *Composite Score* | **15.7 ± 4.1** | **15.4 ± 3.0** | **9.25 ± 5.8** | **9.8 ± 2.9** | **M1 vs M4 ($p=0.027$),**<br>**M2 vs M4 ($p=0.023$)** |

Model 1 = Traditional – Synchronous Model; Model 2 = Traditional – Asynchronous Model; Model 3= Externally Sourced; Model 4= Physician/APP Conducted. DCM= Dilated Cardiomyopathy. Means ± Standard Deviation are shown for descriptive purposes. Statistical differences across groups were assessed using Kruskal-Wallis tests with Dunn's post-hoc tests with Sidak correction for pairwise comparisons. Significance was set at $\alpha=0.05$ . Only significant comparisons ( $p<0.05$ ) are shown.

#### Model 1: The Traditional – Synchronous Model

Six sites used a *Traditional-Synchronous* model. The common theme defining this model included a cardiovascular-specialized GC and cardiologist (synchronous) encounters coordinated within a single, multi-disciplinary clinic visit for the patient. All Model 1 sites provided pre-GT counseling and/or education and family history assessment (Figure 2), with support from an embedded GC (100%, n=6) in addition to an APP (16.7%, n=1) or, in some cases, provided by the cardiologist themself (33%, n=2). Only one site offered genetic evaluation services in-person only, while others offered both telehealth and in-person options (n=5). GCs were the primary team member who returned results to patients, using phone call disclosures across all centers but also offering in-person disclosures (50%, n=3), electronic health record messaging (66%, n=4), and mailed letters (16.7%, n=1). Post-test GC encounters were provided to patients with all types of results (positive, negative, uncertain, and/or incidental findings). Model 1 was the most time-intensive, requiring an estimated cumulative 128 minutes on average per patient to complete all key components (Table 2). This model also provided family-based support post-GT.

#### Model 2: The Traditional – Asynchronous Model

The *Traditional – Asynchronous* model was the most frequently implemented across DCM Consortium sites (n=8), where the cardiology and genetics encounters were separately conducted (asynchronously) with two separate clinic appointments but still followed a historically typical workflow inclusive of genetic counseling both before and after GT (Figure 1). Like Model 1, GCs were identified as a primary team member providing pre-GT counseling and education at a majority (n=7) of sites. Four sites (50%) also listed a cardiologist (including themselves) as a team member providing aspects of this service in addition to GCs. A cardiovascular-specialized GC internal to the institution was a primary team member at all sites that provided results back to patients (100%, n=8) in addition to the cardiologist at two centers (25%) and an APP at one (12.5%). There were several models of post-GT result communication with most Model 2 sites offering multiple approaches including telephone (n=5), in-person (n=5), electronic health record message/letter (n=5), and mailed letter (12.5%, n=1). Model 2 sites provided genetic evaluation components variably (Figure 2), including billing/reimbursement support (87.5%, n=7), reviewing care implications upon receipt of genetic results with the clinical team (75%, n=6), family follow up (75%, n=6), reclassification tracking (75%, n=6), and connection to research (50%, n=4).

#### Model 3: Externally-Sourced Genetics Services

The central feature of the four Model 3 sites was that they referred patients to genetics providers in a department, institution, or company external to the HF and/or cardiology team for coordination of GT (Figure 1). Beyond these components, who, how, and when other aspects of the genetic evaluation were conducted was variable. One Model 3 site had an APP who had specific genetic expertise and/or practice experience with cardiomyopathy to support their program (Table 1), but it was noted that this individual was primarily assigned to hypertrophic cardiomyopathy (HCM) patient navigation. Only two Model 3 sites routinely collected and assessed family history as a part of the genetic evaluation service. Test results were typically communicated back to the patient by the externally referred genetics provider (50%), sometimes in addition to the cardiologist sharing the result themselves (50%) through multiple mechanisms, including telephone (100%), electronic health record communication (75%), or in-person (50%). One Model 3 site only offered post-GT counseling for positive results (pathogenic or likely pathogenic variants) while post-GT counseling was provided for all result types at the other Model 3 sites. Half Model 3 sites provided billing/reimbursement support, reclassification tracking, and connection to research and all reported that they conducted care reviews after receipt of genetic results.

#### Model 4: Physician/APP Conducted Model

Model 4 (n=6 sites) was characterized by a cardiologist with or without the support of an APP partner conducting the genetic evaluation. This was the least time-intensive model, with a cumulative estimate of about 68 minutes required per patient to perform all implemented components (Table 3). Of the two Model 4 sites that did provide pre-GT services, both selected that only “patient education” was provided and this could be done by any clinical team member on the same day the patient was identified in clinic (n=2). Results were provided back to patients by various team members including the cardiologist (66.7%, n=4), an RN (33%, n=2), an APP (16.7%, n=1), or a medical assistant (16.7%, n=1); one site did not specify who disclosed results. **Genetic testing practices**

While implementation of the HFSA indicated components of the genetic evaluation varied widely across and within model groups, all sites across all model types indicated that they routinely conducted GT for DCM in their model (Figure 2). The sample types and test selection varied widely across centers and within practice models (Table 4). Twenty-two PIs stated that GT results changed care varying percentages of patients evaluated: 1-5% (n=2), 5-10% (n=6), 10-20% (n=9), 20-50% (n=4), or 50-100% (n=1). Only two site PIs stated that genetic information never or rarely changed care. When asked if genetic information had been used to make treatment decisions for at least one DCM patient, 87.5% (n=21) selected at least one treatment decision, with ICD implantation (83%; n=20) and cardiac transplant (62.5%; n=15) the most common (Figure 3). Respondents perceived the ability to do GT for HCM was easier to accomplish than for DCM at 42% (n=14) of centers.

**Figure 3.**
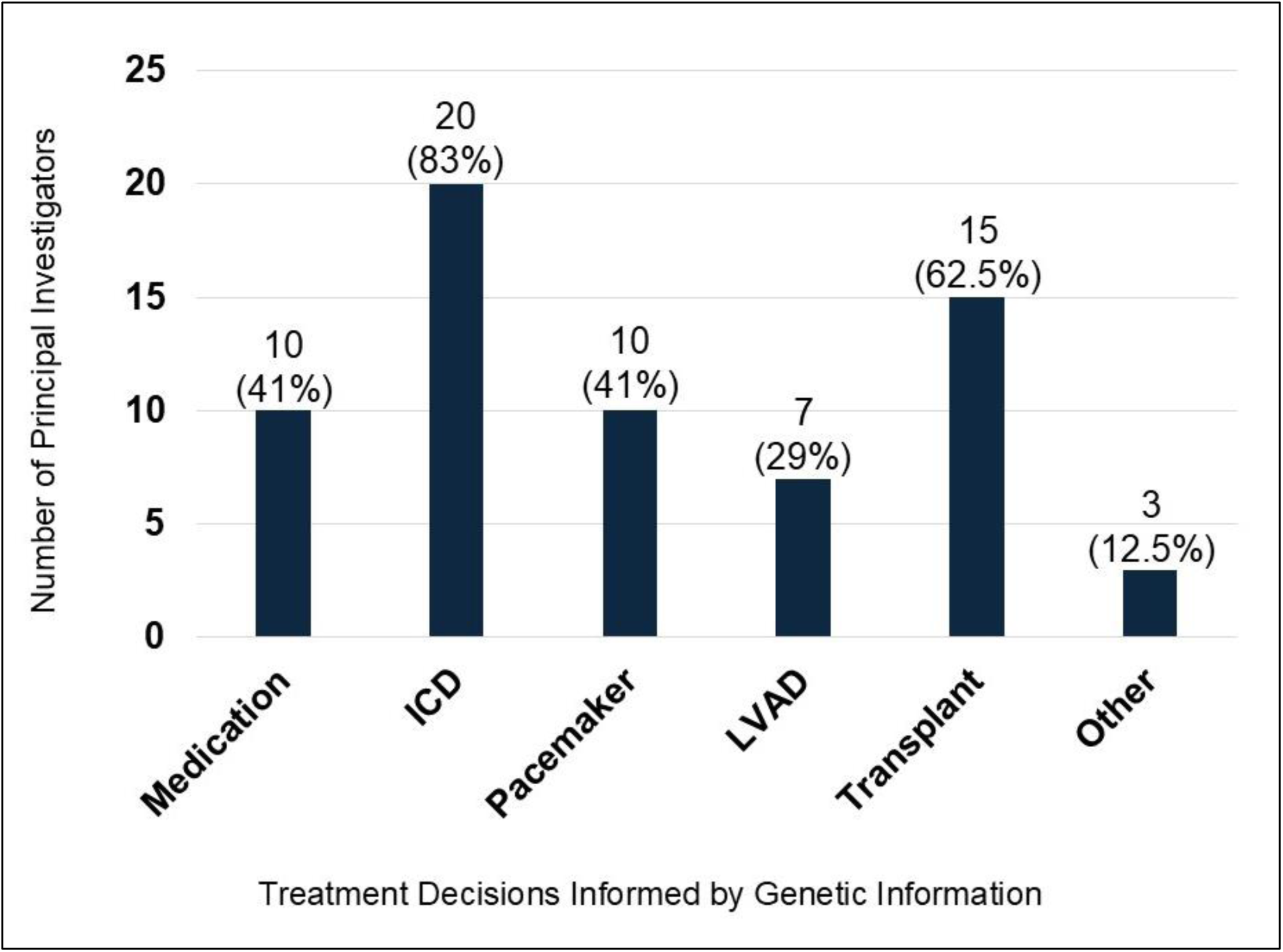
Treatment decisions informed by genetic information for at least one DCM patient by cardiologist site principal investigators of the DCM Consortium. ICD=implantable cardioverter defibrillator; LVAD=left ventricular assist device; Other selections included exercise restrictions (1), screening for sudden death with cardiac monitoring (1), and frequency of follow-up (1).

**Table 4.** Sample collection and genetic testing strategies at DCM Consortium Sites by Model Type.

|  | **Model 1** | **Model 2** | **Model 3** | **Model 4** |
| --- | --- | --- | --- | --- |
| *Sample Type Collected* | n=6<br>(%, n) | n=8<br>(%, n) | n=4<br>(%, n) | n=6<br>(%, n) |
| Blood | 17%, 1 | 0 | 25%, 1 | 17%, 1 |
| Saliva | 0 | 25%, 2 | 0 | 0 |
| Buccal | 33%, 2 | 25%, 2 | 25%, 1 | 33%, 2 |
| Blood or saliva | 17%, 1 | 0 | 25%, 1 | 0 |
| Blood or buccal | 0 | 38%, 3 | 0 | 0 |
| Blood, saliva, or buccal | 33%, 2 | 12%, 1 | 25%, 1 | 50%, 3 |
| *Panel Ordered* |  |  |  |  |
| DCM Targeted | 0 | 12%, 1 | 25%, 1 | 17%, 1 |
| Comprehensive<br>Cardiomyopathy | 33%, 2 | 50%, 4 | 25%, 1 | 66%, 4 |
| Comprehensive<br>Cardiomyopathy and<br>Arrhythmia | 66%, 4 | 38%, 3 | 50%, 2 | 17%, 1 |

Percentages may not add to 100 due to rounding

### Billing and reimbursement practices

GT was billed to insurance at over half (54%, n=13) of sites. Others utilized sponsored GT programs (n=1) or were not sure or did not specify billing practices (n=10). Of the 17 sites (71%) that indicated they routinely supported patients in navigating billing and reimbursement concerns, a majority (82%, n=14) stated that they did not know how often insurance denials for GT were received, one site did not specify, and those that did estimate (n=2) projected that 5-10% of patients experienced denials. Thirteen PIs stated that they did not know how often their patients were responsible for an out-of-pocket cost higher than desired. Of the three sites that provided estimates, it was projected that patients receive high out of pocket costs 10% (n=1) or 20% (n=2) of the time. In contrast, the estimated frequency of patients expressing concern regarding cost of GT ranged widely across centers from 5-75% (mean=27.5%, median=20%) for those providing an estimate (n=10). On a five-point scale of 1 (not at all concerned) to 5 (very concerned), PIs had generally neutral levels of concern (mean=2.8) related to cost of testing. Regarding billing for GC services, 83% said *yes* and 17% said *maybe* improved reimbursement for GCs would improve the ability to add a GC to the HF team.

### Focus Group Discussion

Themes related to facilitators, or enabling factors, and barriers, or hindering factors,^18^ to a genetic evaluation emerged from within-model small group discussions. These themes and exemplary quotes are shown in Table 5. Having access to a GC on the HF clinical team was a consistently named facilitator to navigating the genetic evaluation process. All groups also discussed the supportive impact that authoritative recommendations directing routine implementation of genetic services in DCM could have, creating a foundation for a best practice accreditation framework for DCM care, similar to the current “*center of excellence*” designation available for HCM care, where a genetic evaluation would be a required service available to all qualifying DCM patients to meet care standards for qualifying HF centers.

**Table 5.** Themes that emerged from Focus Group Discussions within Model Type Groups.

| THEME | SUB-THEME | DEMONSTRATIVE QUOTES |
| --- | --- | --- |
| **Facilitators** | Genetic counselors as members of the HF team | *It's just a relief, I think, to have...genetics.... Most of our colleagues are not at all experts in the genetics side of things. So being able to have an expert in genetics, also an advantage. -MD, Traditional-Asynchronous Model* |
|  |  | *Genetic counseling works. Well, patients are satisfied, and they'll travel long distances to see someone who's got the rare condition and understands the gene that they're troubled with, and a positive result that's involving their family. And so there's a lot of positive sides there. -MD, Traditional-Synchronous Model* |
|  |  | *The other problem is... the variants in genes...the what to do about it, you know, and how it relates specifically to this [patient's] phenotypes. I think it's difficult, at least for me, because I don't have enough knowledge to really understand the little nuances of... if that is going to change my management... And that's why, again, today, genetic counseling experience, that discussion with that individual, may help me make that decision which I don't have right now... -MD, Physician/APP Conducted Model* |
|  | Authoritative guidance directing genetic services as standard of care | *And HCM [hypertrophic cardiomyopathy] clinic actually is a little bit streamlined [for] new referrals for HCM. At our center [referrals] are almost always coming from outside the system. And the director for HCM [clinic] actually has it set up that now [a] genetics counselor visit, upfront for pre-test is part of the streamlined package of stuff that you get when you come... -MD, Traditional-Asynchronous Model* |
|  |  | *They did something like that in the PAH [pulmonary arterial hypertension] administration... trying to create PAH centers of excellence... It's kind of like the idea of like cardiovascular genetic center of excellence, right? ...And that was something, because there was a roadmap, and I don't think we have a roadmap.-MD, Physician/APP Conducted Model* |
|  |  | *I know our institution... is more willing to pay for genetic counselors or administrative assistants, if it's part of that "Center of Excellence" tag. -GC, Physician/APP Conducted Model* |
| **Barriers** | Operational challenges to genetic testing | *There is a drop [off] from wanting to do the test to actually getting tested... [it's a] big problem. There are people that will do [a saliva sample] like two or three times.. I think a lot of it is just the systems of testing is just not smooth?-MD, Externally Sourced Model* |
|  |  | *It's hard to even find a variant [in the medical record]... We actually have Epic Genomics...We've got a package in Epic, but we don't even use it because it takes triple the amount of time to put [in] the information...Actually I had a slideshow of what are the 10 ways [you] can find [a genetic test result in Epic]. Five of them is completely invisible, but really kind of the tracking solutions and downstream is the biggest. -MD, Externally Sourced Model* |
|  | Under-recognition of the genetic causes of DCM | *But basically, the doctors just don't see that 30% of the patients actually may have a genetic basis [for their DCM]. That's the problem, that lack of knowledge. When you put it in that perspective, then you would pay attention. But I think it's something that has not sunk in there... And also the other thing is what to do. This is also the other part, you know also. So then what to do with that? It's not really part of the pathway that I think most cardiologists have in their mind. -MD, Physician/APP Conducted Model* |
|  |  | *I've made a lot of way and a lot of education to try to get other cardiologists to do that [genetic testing] independently of me. It actually has worked...because many times I get someone asking me, "Well, how do I do that?" Or "What do I do with this result?" They don't have a structured genetic counseling team in cardiovascular disease which I've been asking for and trying to [get]. -MD, Physician/APP Conducted Model* |
|  | Limited personnel resources | *We have 11 advanced heart failure cardiologists, but only several of us are using [the] genetic counselor. And there are real scalability issues. That if we were to, if the entire group started doing it, I think we'd swamp the resources we have. -MD, Traditional – Asynchronous Model* |
|  |  | *We in fact need more administrative support. Hiring a GC doesn't take care of the problems. We can't accommodate volumes. They don't have infrastructure in place to support those. -MD, Traditional – Asynchronous Model* |
|  |  | *I think the biggest challenge is volume and I think balancing the benefits of pretest counseling with just getting people tested. -GC, Externally Sourced Model* |
|  | Need for new service delivery strategies | *Having the split model [MD + GC clinic and separate GC only clinic], has helped a lot with wait times. That was one of the biggest issues we identified was just that it was getting filled up, and people were waiting a long time a lot of time. -GC, Traditional-Synchronous Model* |
|  |  | *If someone's already seen one of my...heart failure colleagues, I don't need to see them, but a GC needs to see them. And so those are done separately, usually much faster and much more efficiently. -MD, Traditional-Synchronous Model* |
|  |  | *We do a lot of inpatient counseling. But I think it [inpatient counseling] really catches a...more diverse population... Inpatient time is when family members are around. Then, you know, we can actually talk about family screening at the same time... So we definitely try to encourage all the heart failure faculty members, or even any cardiologist who's attending in the hospital. We encourage them to just refer inpatient. -MD, Traditional-Synchronous Model* |
|  | Cost, billing, and reimbursement | *The companies are evolving, the reimbursement is changing. It's hard even for the genetic counselor to keep up sometimes. -MD, Traditional-Asynchronous Model* |
|  |  | *Sometimes I end up seeing [the patient] not because the actual need for seeing the patient [but for billing]. There's a lot of Medicare patients. -MD, Traditional-Synchronous Model* |
|  |  | *It comes down to money. We reorganized our system into a service line model where, instead of the money for procedures, etc, that are done in the hospital, going to the hospital... Everything in cardiology... comes into one pot, and suddenly they have the money to hire a GC. -MD, Traditional-Asynchronous Model* |
|  |  | *...insurance [companies] think of... one year term... budget and expenses. So they don't really care about what happens [in] 5 years. That's why they don't really encourage... preventive medicine, [or] to invest in that, because they only care about the year." -MD, Physician/APP Conducted Model* |

Internal institutional barriers and external factors hindering implementation of DCM genetic services that were discussed included operational challenges, limited personnel resources, including insufficient GC workforce, and an adverse cost and billing landscape with poor reimbursement of genetic services. Lack of scalability of historical models, termed “*Traditional*” herein, were also discussed as a barrier, with participants sharing successes with how they have adapted “traditional” processes for improved efficiency. Model groups also reflected that there was under-recognition of the genetic contributions of DCM among their HF colleagues, which may increase the routine utilization of genetic services into clinical care if better understood.

## DISCUSSION

We have provided a comprehensive assessment of how genetic evaluations for DCM were conducted at 24 academic medical centers, all of whom were engaged in DCM genetic research and have been committed to clinical genetic care for patients with DCM and their families. Using a systematic questionnaire and in-person focus group interviews in follow up to a prior effort,^14^ four different practice models for genetic care for patients with DCM were identified. While genetic testing was a universal component of all models, other model characteristics varied greatly, as did the time, effort and expertise of the personnel needed to conduct genetic evaluations.

This assessment of care models has provided practical insights for others who are developing cardiomyopathy genetic evaluation programs and seek to elevate their clinical service beyond “*just the test*.” Genetic evaluation of cardiomyopathy is informed by practice guidelines from the Heart Failure Society of America (HFSA)^5^ and American College of Medical Genetics and Genomics (ACMG),^3–6^ where a genetic evaluation is defined as “*a systematic approach that includes a comprehensive family history, phenotypic evaluation of the proband and at-risk family members, genetic counseling, genetic testing, if indicated, with pre- and post-test counseling, and guidance as indicated for specific drug and/or device or other specific therapeutic interventions*.” We emphasize that genetic testing is a minimum necessary component but alone does not define the broader service of a genetic evaluation, which extends the clinical impact of genetic information to patient care and family risk-management. Until genetic evaluations for DCM are able to be implemented *routinely for all* qualifying patients, patients and families will suffer from the missed opportunities that precision care for DCM now offers.^1^

While all models consistently implemented the minimum component of genetic testing, the key differentiating characteristic across models centered on whether a clinical genetics team member, typically a GC but in some cases an APP with genetics clinical focus, was accessible to partner with and/or support physicians with genetic evaluation components, and, if so, if they were housed internal or external to the HF team or institution. Models 1 and 2 utilized a “traditional” model inclusive of pre- and post-test genetic counseling services provided by cardiovascular-specialized GCs. This workflow has been historically considered a gold standard approach and rated as significantly more acceptable^15^ in the current report. Access to a GC has repeatedly been identified as a facilitator to implementing clinical genetics services for cardiovascular disease.^14^ While the historical embedded model (termed here as a “*Traditional-Synchronous/Model 1*”) was shown to provide comprehensive services, such a standard may sacrifice a more equitable provision of services due to the lack of scalability. Notably, the traditional model was the most time- and resource-intensive approach, requiring nearly double the time compared to the other models reported herein. Alternative models of service delivery are actively being piloted internationally in multiple specialty areas including HF clinics.^20,21^ In order to fully realize the potential of new approaches, improved billing and reimbursement procedures are needed in the US, which has been a consistent major barrier to genetics services.^22,23^ New billing codes for GC services are available and the community has continued to appeal to CMS^23^ to acknowledge GCs as billable providers.^3,5,24–26^ Although lack of GC support was a barrier theme emerging from the discussions of all model groups, GC-integrated programs were found to generate a positive downstream revenue,^27^ underscoring the fiscal value of building genetics expertise into cardiovascular programs.

The broad-based discussions brought forward additional insight relevant to considering the implementation of genetic care. In an open text item, a PI suggested that cardiologists need to “*lower the bar to refer DCM patients for genetic evaluation*,” a sentiment aligning with the barrier of under-recognition of genetic contributions to DCM amongst the HF clinical community (Table 5). Indeed, all qualifying patients with DCM and their family members, regardless of severity of their personal or family history at diagnosis, should have the opportunity to seek precision care guided by the remarkable and emerging benefits that a genetic diagnosis of cardiomyopathy can now facilitate. However, as we lower the bar to ensure all DCM patients have access, we must also take care to raise the bar for clinical care standards as novel, scalable adaptations to current models are developed.

It was also speculated in an open response item that “*The oncology and obstetrics models of genetic evaluations are much further ahead because genomic evaluation is part of their clinical management processes and not taken out of the practice pathway.*” Indeed, there are authoritative and time-specific recommendations for GT and genetic counseling from organizations such as the American College of Obstetrics and Gynecology^28–31^ and the National Comprehensive Cancer Network,^32^ driven by scientific advancements that established a direct impact of genetic information on immediate treatment and care decisions in these areas.

Underlying the remarkable uptake of cancer genetics is the direct impact of cancer tissue somatic genetic signatures on chemotherapeutic treatment decisions. Within the DCM and HF care paradigm germline, the evidence base of genetic data has not been produced to directly inform treatment algorithms. Such studies are needed for HF patients with DCM, as underscored in the results reported here ∼88% of PIs, who are HF/TX cardiologists, are already actively using genetic information to inform clinical decisions based on conventional wisdom, expert consensus, and clinical experience with genetic information.

The highly variable models of genetic care shown in this study suggest that a more sustained and substantial evaluation coupled with innovation is needed for DCM care models, as DCM is the most common cardiomyopathy and accounts for a sizable proportion of HF from reduced ejection fraction (HFrEF). Patients with HFrEF now commonly receive care within HF clinics. Interestingly, 42% of DCM Consortium site PIs reported that genetic evaluations are more routinely accomplished for patients with HCM than for DCM. Notably HCM clinics at centers meeting a HCM Center of Excellence designation require genetics services. In contrast, defining the genetic etiology of DCM in HF clinics has not been a clinical priority.^33^ We suggest one insight derived from this analysis is that developing and establishing an authoritative best practice framework inclusive of clinical genetics practice guidance and accreditation mechanisms for the care for all cardiomyopathies is needed. If achieved, such a framework may drive awareness and implementation of genetic medicine for all HF patients with DCM and other related cardiomyopathies (e.g., ARVC, HCM, etc), as applicable, seen in HF or other relevant clinics.

### Limitations

This assessment only represents the experiences and perspectives of those of the DCM Consortium, all of whom are members of advanced HF/TX programs at academic medical centers and therefore is not representative of practices beyond academia or of all HF clinical team members. The time required to implement evaluation components were estimates by PIs completing the needs assessment survey, and in some cases were estimates for service components that they may not have routinely provided. In addition, not all DCM Consortium site PIs or GCs or APPs from all sites were able to attend the Summer Scientific Symposium. Given time constraints, not all focus groups were able to discuss all questions. Focus group facilitators were GCs, a key genetics professional group of this study, however a COREQ checklist (Supplemental Material), has provided additional transparency on positionality of the facilitators. Despite limitations, the PI responses reported herein provide a broad and clinically relevant set of perspectives, as DCM Consortium PIs had expertise to critically reflect on the patterns and needs of the practice of genetics care in HF as shared through qualitative analysis.

## CONCLUSIONS

Current care models for the provision of genetic services across the sites of the DCM Consortium are highly variable. To address the need for genetic evaluation to be equitably accessible to all DCM patients, identifying practice standards to implement components of a DCM genetic evaluation that can be adapted and applied across highly variable institutional environments may yield more scalable clinical genetics care models.

## Data Availability

The data can be made available upon reasonable request.

## Acknowledgements

None.

## SUPPLEMENTAL MATERIAL

Appendix 1. Survey instrument

Appendix 2. Focus group interview guide

Appendix 3. COREQ Checklist

Appendix 4. Codebook and counts

## REFERENCES

1. Parikh VN, Day SM, Lakdawala NK, Adler ED, Olivotto I, Seidman CE, Ho CY. Advances in the study and treatment of genetic cardiomyopathies. Cell. 2025;188:901–918. doi: 10.1016/j.cell.2025.01.011

2. Landstrom AP, Ferguson JF, James CA, Key KV, Lanfear D, Natarajan P, Rasmussen-Torvik LJ, Reza N, Roden DM, Tsao PS, et al. Genetic and Genomic Testing in Cardiovascular Disease: A Policy Statement From the American Heart Association. Circulation. 2025;152:e474–e489. doi: 10.1161/CIR.0000000000001385

3. Musunuru K, Hershberger RE, Day SM, Klinedinst NJ, Landstrom AP, Parikh VN, Prakash S, Semsarian C, Sturm AC, American Heart Association Council on G, et al. Genetic Testing for Inherited Cardiovascular Diseases: A Scientific Statement From the American Heart Association. Circ Genom Precis Med. 2020;13:e000067. doi: 10.1161/HCG.0000000000000067

4. Wilde AAM, Semsarian C, Marquez MF, Sepehri Shamloo A, Ackerman MJ, Ashley EA, Sternick EB, Barajas-Martinez H, Behr ER, Bezzina CR, et al. European Heart Rhythm Association (EHRA)/Heart Rhythm Society (HRS)/Asia Pacific Heart Rhythm Society (APHRS)/Latin American Heart Rhythm Society (LAHRS) Expert Consensus Statement on the State of Genetic Testing for Cardiac Diseases. Heart Rhythm. 2022;19:e1–e60. doi: 10.1016/j.hrthm.2022.03.1225

5. Hershberger RE, Givertz MM, Ho CY, Judge DP, Kantor PF, McBride KL, Morales A, Taylor MRG, Vatta M, Ware SM. Genetic Evaluation of Cardiomyopathy-A Heart Failure Society of America Practice Guideline. J Card Fail. 2018;24:281–302. doi: 10.1016/j.cardfail.2018.03.004

6. Hershberger RE, Givertz MM, Ho CY, Judge DP, Kantor PF, McBride KL, Morales A, Taylor MRG, Vatta M, Ware SM, et al. Genetic evaluation of cardiomyopathy: a clinical practice resource of the American College of Medical Genetics and Genomics (ACMG). Genet Med. 2018;20:899–909. doi: 10.1038/s41436-018-0039-z

7. Morales A, Moretz C, Ren S, Smith E, Callis TE, Hall T, Hatchell KE, Nussbaum RL, Regalado E, Rojahn S, et al. Real-World Genetic Testing Utilization Among Patients With Cardiomyopathy. Circ Genom Precis Med. 2024;17:e004028. doi: 10.1161/CIRCGEN.122.004028

8. Hellwig LD, Banaag A, Olsen C, Turner C, Haigney M, Koehlmoos T. A health systems assessment of genetic counseling in cardiovascular care in a large health system: Adherence to genetics recommendations in the Military Health System. J Genet Couns. 2024;33:888–896. doi: 10.1002/jgc4.1791

9. Longoni M, Bhasin K, Ward A, Lee D, Nisson M, Bhatt S, Rodriguez F, Dash R. Real-world utilization of guideline-directed genetic testing in inherited cardiovascular diseases. Front Cardiovasc Med. 2023;10:1272433. doi: 10.3389/fcvm.2023.1272433

10. Bui QM, Morales A, Adler ED, Urey MA, Sinha A, Wang J, Hong KN, Ramsis M. Real-world Genetic Testing Practices in Cardiomyopathy: National Insights From 1.7 Million Patients. J Card Fail. 2026. doi: 10.1016/j.cardfail.2025.11.496

11. Morales A, Goehringer J, McDonald PL, Sanoudou D. Implementation of genetic testing for heritable cardiac conditions: A scoping review. Genet Med Open. 2025;3:103441. doi: 10.1016/j.gimo.2025.103441

12. Tang WHW, Bui QM, Cirino AL, Dellefave-Castillo L, Floyd BJ, Guerchicoff A, Guerchicoff M, A VK, Knowles JW, Lafayette K, et al. Cardiologists’ Perceptions of Cardiogenetic Testing and Management. JACC Adv. 2025;4:101910. doi: 10.1016/j.jacadv.2025.101910

13. Kinnamon DD, Morales A, Bowen DJ, Burke W, Hershberger RE, Consortium* DCM. Toward Genetics-Driven Early Intervention in Dilated Cardiomyopathy: Design and Implementation of the DCM Precision Medicine Study. Circ Cardiovasc Genet. 2017;10. doi: 10.1161/CIRCGENETICS.117.001826

14. Jordan E, Ni H, Parker P, Kinnamon DD, Owens A, Lowes B, Shenoy C, Martin CM, Judge DP, Fishbein DP, et al. Implementing Precision Medicine for Dilated Cardiomyopathy: Insights From the DCM Consortium. Circ Genom Precis Med. 2025;18:e005078. doi: 10.1161/CIRCGEN.125.005078

15. Weiner BJ, Lewis CC, Stanick C, Powell BJ, Dorsey CN, Clary AS, Boynton MH, Halko H. Psychometric assessment of three newly developed implementation outcome measures. Implement Sci. 2017;12:108. doi: 10.1186/s13012-017-0635-3

16. Proctor E, Silmere H, Raghavan R, Hovmand P, Aarons G, Bunger A, Griffey R, Hensley M. Outcomes for implementation research: conceptual distinctions, measurement challenges, and research agenda. Adm Policy Ment Health. 2011;38:65–76. doi: 10.1007/s10488-010-0319-7

17. Tong A, Sainsbury P, Craig J. Consolidated criteria for reporting qualitative research (COREQ): a 32-item checklist for interviews and focus groups. Int J Qual Health Care. 2007;19:349–357. doi: 10.1093/intqhc/mzm042

18. Damschroder LJ, Aron DC, Keith RE, Kirsh SR, Alexander JA, Lowery JC. Fostering implementation of health services research findings into practice: a consolidated framework for advancing implementation science. Implement Sci. 2009;4:50. doi: 10.1186/1748-5908-4-50

19. Glaser BG, Strauss A. A discovery of grounded theory: Strategies for qualitative research. Chicago: Aldine; 1967.

20. Mackley MP, Richer J, Guerin A, Caluseriu O, Armstrong L, Blood KA, Bernier F, Boswell-Patterson C, Chard M, Costain G, et al. Mainstreaming of clinical genetic testing: A conceptual framework. Genet Med. 2025;27:101465. doi: 10.1016/j.gim.2025.101465

21. Mohananey A, Tseng AS, Julakanti RR, Gonzalez-Bonilla HM, Kruisselbrink T, Prochnow C, Rodman S, Lin G, Redfield MM, Rosenbaum AN, et al. An intervention strategy to improve genetic testing for dilated cardiomyopathy in a heart failure clinic. Genet Med. 2023;25:100341. doi: 10.1016/j.gim.2022.11.009

22. Landstrom AP, Ferguson JF, James CA, Key KV, Lanfear D, Natarajan P, Rasmussen-Torvik LJ, Reza N, Roden DM, Tsao PS, et al. Genetic and Genomic Testing in Cardiovascular Disease: A Policy Statement From the American Heart Association. Circulation. 2025. doi: 10.1161/CIR.0000000000001385

23. Riordan S, Richardson J, Zierhut H, Goodnight BL, Sieling FH, Black CM, Moore RA. Medicare beneficiary barriers to genetic counselor services: Implications for patient policy, decision-making, and care. J Genet Couns. 2024;33:262–268. doi: 10.1002/jgc4.1729

24. Sturm AC, Knowles JW, Gidding SS, Ahmad ZS, Ahmed CD, Ballantyne CM, Baum SJ, Bourbon M, Carrie A, Cuchel M, et al. Clinical Genetic Testing for Familial Hypercholesterolemia: JACC Scientific Expert Panel. J Am Coll Cardiol. 2018;72:662–680. doi: 10.1016/j.jacc.2018.05.044

25. Isselbacher EM, Preventza O, Hamilton Black J, 3rd, Augoustides JG, Beck AW, Bolen MA, Braverman AC, Bray BE, Brown-Zimmerman MM, Chen EP, et al. 2022 ACC/AHA Guideline for the Diagnosis and Management of Aortic Disease: A Report of the American Heart Association/American College of Cardiology Joint Committee on Clinical Practice Guidelines. Circulation. 2022;146:e334–e482. doi: 10.1161/CIR.0000000000001106

26. Wilde AAM, Semsarian C, Marquez MF, Sepehri Shamloo A, Ackerman MJ, Ashley EA, Sternick Eduardo B, Barajas-Martinez H, Behr ER, Bezzina CR, et al. European Heart Rhythm Association (EHRA)/Heart Rhythm Society (HRS)/Asia Pacific Heart Rhythm Society (APHRS)/Latin American Heart Rhythm Society (LAHRS) Expert Consensus Statement on the state of genetic testing for cardiac diseases. J Arrhythm. 2022;38:491–553. doi: 10.1002/joa3.12717

27. Olson M, Anderson J, Knapke S, Kushner A, Martin L, Statile C, Shikany A, Miller EM. Cardiac genetic counseling services: Exploring downstream revenue in a pediatric medical center. J Genet Couns. 2025;34:e1984. doi: 10.1002/jgc4.1984

28. American College of O, Gynecologists’ Committee on Practice B-O, Committee on G, Society for Maternal-Fetal M. Screening for Fetal Chromosomal Abnormalities: ACOG Practice Bulletin, Number 226. Obstet Gynecol. 2020;136:e48–e69. doi: 10.1097/AOG.0000000000004084

29. ACOG Committee Opinion No. 486: Update on carrier screening for cystic fibrosis. Obstet Gynecol. 2011;117:1028–1031. doi: 10.1097/AOG.0b013e31821922c2

30. ACOG Committee Opinion No. 469: Carrier screening for fragile X syndrome. Obstet Gynecol. 2010;116:1008–1010. doi: 10.1097/AOG.0b013e3181fae884

31. ACOG Committee Opinion No. 442: Preconception and prenatal carrier screening for genetic diseases in individuals of Eastern European Jewish descent. Obstet Gynecol. 2009;114:950. doi: 10.1097/AOG.0b013e3181bd12f4

32. Network NCC. Genetic/Familial High-Risk Assessment: Breast, Ovarian, Pancreatic, and Prostate. https://www.nccn.org/guidelines/category_2. 2025. Accessed 14 November.

33. Tayal U, Prasad S, Cook SA. Genetics and genomics of dilated cardiomyopathy and systolic heart failure. Genome Med. 2017;9:20. doi: 10.1186/s13073-017-0410-8

